# Beyond the Bowel: Novel Comorbidity Patterns in Inflammatory Bowel Disease from the All of Us Research Program

**DOI:** 10.1101/2025.06.01.25328740

**Authors:** Soham C. Sudhakaran, Matthew T. Wayland, Yogesh Purushothaman, Scott B. Minchenberg, Kanwal Bains, Sudhakaran Prabakaran

## Abstract

**Background:** Inflammatory bowel disease (IBD) manifests systemically, yet most comorbidity studies rely on predominantly European populations. The All of Us Research Program enables investigation across demographically diverse groups.

**Methods:** We matched 5,094 IBD patients 1:4 with controls by age, gender, and race, analyzing comorbidities using logistic regression with Mantel-Haenszel adjustment. Multiple testing correction used false discovery rate (FDR) with significance thresholds of OR >1.5 or <0.5 and FDR <0.05.

**Results:** Our cohort included 29.2% non-White participants versus 10-15% in traditional studies. We identified 22 significant associations across seven organ systems. Three novel discoveries included delayed postmyocardial infarction pericarditis (adjusted OR = 4.80), contact dermatitis (adjusted OR = 1.84), and carotid artery aneurysm (adjusted OR = 2.21). Other significant associations included drug-induced lupus (adjusted OR = 4.32), autoimmune hepatitis (adjusted OR = 2.43), and restricting-type eating disorders (adjusted OR = 4.00). IBD patients showed decreased obesity-related conditions.

**Conclusion:** This demographically diverse study discovered novel IBD comorbidities and confirmed established associations across racial groups. Findings support reconceptualizing IBD as a multisystem disorder requiring comprehensive management and demonstrate the importance of diverse research populations.

## Introduction

### The Rising Burden of Inflammatory Bowel Disease

Inflammatory bowel disease (IBD), comprising Crohn’s disease (CD) and ulcerative colitis (UC), affects approximately 4.9 million people globally, with prevalence increasing from 3.3 million in 1990 (Wang et al. 2023; Hracs et al. 2025). In the United States, IBD affects 0.7% of the population and imposes substantial economic burden, with patients incurring three times higher healthcare costs than controls ($22,987 vs. $6,956 annually) (Hudesman et al. 2020). Beyond gastrointestinal symptoms, IBD significantly impacts quality of life and contributes to 1.62 million disability-adjusted life years globally (Zhou et al. 2023).

### IBD Comorbidities: Current Understanding and Limitations

IBD comorbidities—secondary health conditions beyond the primary gastrointestinal diagnosis— significantly complicate disease management and worsen clinical outcomes. Large-scale studies have documented extensive comorbidity burden, with 78% of IBD patients having at least one comorbidity, particularly rheumatologic, dermatologic, and autoimmune conditions (Ghersin et al. 2020). Recent neuropsychiatric research using All of Us data revealed that 54% of patients with psychiatric diagnoses have multiple concurrent conditions, highlighting complex multimorbidity patterns (Barr, Bigdeli, and Meyers 2022).

Despite these advances, significant knowledge gaps persist. Most studies focus on predominantly White European populations, underrepresenting racial minorities and diverse demographics. Additionally, few studies have comprehensively examined rare cardiovascular, neurological, and metabolic comorbidities, creating gaps in understanding IBD’s true systemic burden (Argollo et al. 2019).

### Addressing Demographic Limitations in IBD Research

A critical limitation of existing IBD comorbidity research is the demographic homogeneity of study populations. Major European cohorts, including the Swiss IBD Cohort Study and Dutch IBD Biobank, demonstrate excellent research quality but reflect predominantly Caucasian populations with minimal ethnic diversity (Pittet et al. 2019; Spekhorst et al. 2017). Similarly, large North American IBD studies show striking underrepresentation of racial minorities, with cohorts typically comprising 80-90% White participants despite the growing IBD burden among diverse populations. The SPARC IBD study reports 80.5% White participants, while the TARGET-IBD registry includes 86.6% Caucasian participants, despite the U.S. population being only 60% non-Hispanic White (Raffals et al. 2022; Click et al. 2021). This demographic limitation is particularly concerning given that IBD incidence increased 134% in racial minorities versus 39% in Whites between 1970-2010 (Aniwan et al. 2019), and that genome-wide association studies in African Americans have identified population-specific disease loci completely absent from European cohorts (Huang et al. 2015). The All of Us Research Program addresses these critical gaps by achieving unprecedented diversity in IBD research, with 70% White participants compared to 85-90% in traditional cohorts, and 77% of overall participants from historically underrepresented groups (Ramirez et al. 2022; All of Us Research Program Investigators et al. 2019). This demographic representativeness is essential for identifying population-specific comorbidity patterns and ensuring research findings are generalizable across the increasingly diverse populations affected by IBD.

### Rationale for Comprehensive Investigation

To address these limitations, we employed a systematic approach targeting IBD’s full spectrum of potential manifestations across four theoretical frameworks: (1) shared inflammatory pathways involving IL-17/IL-23/IL-12TNF-α networks; (2) epithelial barrier dysfunction extending beyond the intestinal tract; (3) systemic inflammation consequences affecting cardiovascular and metabolic systems; and (4) gut-organ axis dysfunction involving bidirectional microbiome-organ communication. Our initial comorbidity framework utilized broad clinical categories commonly referenced in medical literature (Supplementary Table 1). To ensure high case specificity, we implemented a validated two-step case definition requiring both IBD diagnostic codes and evidence of prescription for IBD-specific medications (Supplementary Table 2), following established best practices for population-based IBD research. The All of Us Research Program returned results using the standardized OMOP Common Data Model, which employs granular diagnostic classifications, resulting in identification of specific clinical entities rather than general disease categories. For example, rather than the broad category of “kidney stones,” the database identified specific compositions such as calcium oxalate and uric acid calculi. This data-driven approach also revealed associations with conditions not initially hypothesized, including pregnancy-related complications and specific cardiovascular phenomena, demonstrating the value of comprehensive exploration in identifying novel disease associations.

### Study Aim

This study utilizes the All of Us Research Program’s large, diverse dataset to systematically investigate comorbidity associations with IBD. Using a validated case definition combining diagnostic codes with IBD-specific medication requirements, we analyzed the complete spectrum of conditions identified through OMOP coding (Supplementary Table 1) to characterize the strength of association between IBD and dermatological, cardiovascular, autoimmune, and other systemic conditions. Our approach prioritizes rare and understudied comorbidities that have received limited attention in previous literature, thereby informing future surveillance and therapeutic strategies for diverse IBD populations.

## Methods

### Study Design and Data Source

This case-control study utilized data from the All of Us Research Program, a nationwide precision medicine initiative led by the National Institutes of Health (NIH) established in 2016 (All of Us Research Program Investigators et al. 2019). The program aims to recruit a diverse cohort of at least one million individuals in the United States to accelerate biomedical research and improve individualized healthcare. The All of Us Research Program places special emphasis on including participants from groups historically underrepresented in biomedical research, making it particularly valuable for studying health disparities and diverse population characteristics.

For this analysis, we accessed the Registered Tier dataset through the Researcher Workbench platform, which provides individual-level data including electronic health records (EHRs), survey responses, and physical measurements. All data were harmonized to the Observational Medical Outcomes Partnership (OMOP) Common Data Model version 5.2, which standardizes clinical terminology and facilitates consistent analysis across various data sources. The OMOP model organizes patient information into standardized tables for conditions, procedures, measurements, observations, and other clinical domains.

The analysis period spanned from October 2024 to February 2025. All research activities were conducted within the secure cloud-based Researcher Workbench environment in compliance with the All of Us Data (version 7) User Code of Conduct. This study was approved by our institutional review board and was deemed non-human subjects research due to the use of de-identified data provided through the All of Us Research Program.

### Study Population

#### Dataset Overview

At the time of analysis, the All of Us Research Program dataset contained 413,457 participants with electronic health record (EHR) data available for analysis. From this population, we identified participants with IBD and related conditions using standardized diagnostic criteria.

#### Case Definition

IBD cases were identified from the All of Us EHR data using standardized OMOP concept codes in the condition_occurrence table. Specifically, participants with at least one occurrence of any of the following diagnostic codes were classified as having IBD:

- 4074815 (Inflammatory bowel disease)
- 64766004 (Crohn’s disease)
- 34000006 (Ulcerative colitis)

The comprehensive IBD population breakdown was as follows:

- **Total IBD cases (any IBD diagnosis)**: 5,094 participants
- **Ulcerative colitis**: 2,868 participants
- **Crohn’s disease**: 2,997 participants
- **General IBD (unspecified)**: 2,236 participants

*Note: These numbers reflect overlapping diagnoses, as some participants had multiple IBD-related diagnostic codes recorded in their medical records*.

For the purpose of this analysis, all participants with any IBD-related diagnostic code were classified as having IBD and analyzed as a single cohort. When participants had multiple specific diagnoses (e.g., both Crohn’s disease and ulcerative colitis codes), they were included in the general IBD category rather than being assigned to specific subtypes. This approach was chosen to: (1) ensure adequate statistical power for rare comorbidity detection, (2) avoid misclassification issues when clinical distinction between subtypes may be unclear, and (3) capture the full spectrum of IBD-related systemic manifestations regardless of subtype.

No additional temporal criteria or requirements for multiple occurrences were applied. This inclusive approach allowed us to capture all participants with an IBD diagnosis in the All of Us dataset. The index date for cases was defined as the date of the first recorded IBD diagnosis.

We excluded participants with missing demographic information required for matching. After applying these criteria, our final analytical sample included 5,094 IBD cases for analysis. We did not conduct separate subgroup analyses for Crohn’s disease and ulcerative colitis patients; all IBD patients were analyzed as a single cohort to ensure adequate statistical power for rare comorbidity detection.

#### Control Selection

The control cohort comprised participants without any recorded instances of the aforementioned IBD-related concept codes (4074815, 64766004, 34000006) in their condition_occurrence records. For each confirmed IBD case, we selected four controls using a 1:4 matching ratio to enhance statistical power while maintaining computational efficiency.

The Cohort Matcher tool, an integrated component of the All of Us Researcher Workbench platform, was employed to implement the matching algorithm. Controls were matched to cases based on three demographic variables:

- Age (within ±1 year)
- Gender (exact match)
- Race (exact match)

This approach was designed to minimize confounding by these fundamental demographic factors. The matching process was performed without replacement, meaning each control participant could be matched to only one case. When multiple potential controls met the matching criteria for a case, selections were made randomly from the pool of eligible controls. For cases where fewer than four matching controls were available, all available matching controls were included, and the reduced matching ratio was accounted for in the statistical analysis.

Although our study design included matching controls to cases on age, gender, and race, analysis of the final study population revealed significant differences in these demographic variables between groups (**Table 1**). This discrepancy occurred because the complexity of demographic data labels and subcategories in the All of Us Research Program dataset made achieving perfect 1:1 matching challenging for all participants. We prioritized including all eligible IBD cases to maximize statistical power for detecting rare comorbidities, which occasionally required compromises in the strictness of matching criteria. To account for these demographic differences, we performed additional statistical adjustments using the Mantel-Haenszel method as described in our statistical analysis section, which controlled for these demographic variables in our calculation of adjusted odds ratios.

**Table 1:** Demographic and Clinical Characteristics of IBD Cases and Controls. All demographic comparisons were performed between IBD cases (n=5,094) and matched controls (n=20,376) using a 1:4 case-control ratio. Percentages are calculated within each group. For “Race/Ethnicity,” “Gender,” “Annual household income,” “Educational attainment,” and “Health insurance” categories, the overall p-value reflects the statistical significance of distribution differences between cases and controls across all subcategories. Missing or unspecified data were included in statistical analyses but do not necessarily indicate significant differences in these specific subcategories.

| Characteristic | Cases, number (%)<br>(n=5,094) | Controls, number (%)<br>(n=20,376) | p-value* |
| --- | --- | --- | --- |
| **Race/Ethnicity** |  |  |  |
| White | 3,608 (70.8) | 11,384 (55.9) | < 0.001 |
| Black or African American | 624 (12.2) | 4,405 (21.6) |  |
| None Indicated | 454 (8.9) | 2,735 (13.4) |  |
| More than one population | 101 (2.0) | 361 (1.8) |  |
| Asian | 68 (1.3) | 363 (1.8) |  |
| Other/Not specified | 239 (4.7) | 1,128 (5.5) |  |
| **Gender** |  |  |  |
| Female | 3,091 (60.7) | 11,760 (57.7) | < 0.001 |
| Male | 1,846 (36.2) | 7,963 (39.1) |  |
| Other/Not specified | 157 (3.1) | 653 (3.2) |  |
| **Age (years)** |  |  |  |
| Average | 59.0 | 62.6 |  |
| Standard Deviation | 16.6 | 15.0 |  |
| **Annual household income (\$)** | | | |
| ≥ 75,000 | 1,683 (33.0) | 5,439 (26.7) | < 0.001 |
| 35,000-74,999 | 1,397 (27.4) | 5,412 (26.6) |  |
| < 35,000 | 1,057 (20.7) | 4,867 (23.9) |  |
| Prefer not to answer/Skip | 957 (18.8) | 4,658 (22.9) |  |
| **Educational attainment** |  |  |  |
| College graduate or advanced degree | 1,648 (32.4) | 8,155 (40.0) | < 0.001 |
| Some college | 1,003 (19.7) | 5,790 (28.4) |  |
| High school graduate or GED | 559 (11.0) | 3,964 (19.5) |  |
| Less than high school | 158 (3.1) | 1,779 (8.7) |  |
| Not specified/Missing | 1,726 (33.9) | 688 (3.4) |  |
| **Health insurance** |  |  |  |
| Yes | 3,338 (65.5) | 19,063 (93.6) | < 0.001 |
| No | 48 (0.9) | 592 (2.9) |  |
| Not specified/Missing | 1,708 (33.5) | 721 (3.5) |  |

\*p-values based on Pearson $\chi^2$ or Fisher's exact test, as appropriate. P-values < 0.05 were considered statistically significant.

### Additional exclusion criteria for controls included

- Participants with missing demographic information required for matching
- Participants with incomplete data for the analyzed comorbid conditions

Controls were assigned the same index date as their matched cases to ensure comparable observation periods. The final study population included 5,094 IBD cases and 20,376 matched controls.

#### Demographic and Clinical Data

Demographic characteristics, including race/ethnicity, gender, age, annual household income, educational attainment, and health insurance status, were extracted from both the structured EHR data and participant survey responses in the All of Us dataset. In cases where discrepancies existed between EHR and survey data, we prioritized self-reported information from surveys to better reflect participants’ self-identification.

Race/ethnicity was categorized as White, Black or African American, Asian, More than one population, Other/Not specified, and None Indicated, based on standard All of Us classifications. Age was calculated at the index date for both cases and controls. Annual household income was stratified into four categories: ≥$75,000, $35,000-$74,999, <$35,000 and Prefer not to answer/Skip. Educational attainment was classified as College graduate or advanced degree, Some college, High school graduate or GED, Less than high school, and Not specified/Missing. Health insurance status was recorded as Yes, No, or Not specified/Missing.

#### Statistical Analysis of Demographic Characteristics

Demographic characteristics were compared between IBD cases and matched controls using appropriate statistical tests. For categorical variables (race/ethnicity, gender, annual household income, educational attainment, and health insurance status), we employed Pearson χ² tests when expected cell counts were ≥5, and Fisher’s exact tests when expected cell counts were <5. For continuous variables such as age, independent samples t-tests were used to compare means between groups. Statistical significance was set at p < 0.05 for all demographic comparisons. The results of these demographic comparisons are presented in **Table 1**.

### Comorbidity Identification

#### Comorbidity Selection Strategy

Comorbid conditions for analysis were selected based on a comprehensive, predefined list compiled through systematic literature review and clinical expertise to include conditions with potential mechanistic relationships or epidemiological associations with IBD. The selection process was designed to capture both well-established IBD-associated conditions and novel potential associations across multiple organ systems.

Our initial screening identified a broad range of potential comorbidities organized into major categories:

- **Dermatological conditions** including acne, hidradenitis suppurativa, alopecia areata, contact dermatitis, dermatomyositis, eczema, lichen planus, localized scleroderma, psoriasis, seborrheic dermatitis, and vitiligo.
- **Cardiovascular and vascular conditions** including pericarditis, myocarditis, hypertensive disorders, and various arterial conditions.
- **Autoimmune and rheumatologic conditions** including diabetes mellitus (types 1 and 2), autoimmune hepatitis, Sjögren’s syndrome, and rheumatoid arthritis.
- **Renal and urological conditions** including end-stage renal disease, kidney stones, nephritis, and various renal complications.
- **Neurological and psychiatric conditions** including seizure disorders, anxiety, depression, and eating disorders.
- **Gastrointestinal conditions** beyond IBD, including various complications and related disorders.
- **Respiratory conditions** including interstitial lung disease and lung cancer.

The complete list of investigated comorbidities is provided in **Supplementary Table 1**, which details both the proposed conditions for investigation and the specific conditions returned by the All of Us Research Program query results.

### Data Extraction and Processing

We created comprehensive SQL queries in the All of Us Researcher Workbench to extract condition occurrence records for both cases and controls. Each comorbid condition was identified using corresponding OMOP standard concept codes in the condition_occurrence table, ensuring standardized and reproducible case definitions.

For each participant, we identified all diagnosed conditions recorded within the observation period, which extended from the earliest available EHR entry to the end of the data collection period (February 2025). The data extraction process involved several key steps:

**1. Query Construction:**

- OMOP concept codes were systematically mapped to identify cases, controls, and all specified comorbidities
- SQL queries were developed and validated within the All of Us Researcher Workbench environment
- Demographic and clinical variables were extracted for all participants using standardized data dictionaries
**2. Comorbidity Prevalence Assessment:**

- Among the 413,457 total participants in the dataset, **186,581 participants** (45.1%) had at least one of the investigated comorbidities
- Of the 5,094 IBD cases, **3,469 participants** (68.1%) had at least one comorbidity
- This high comorbidity prevalence in IBD cases compared to the general dataset population provided the foundation for our comparative analysis
**3. Data Integration and Quality Control:**

- Case and control data were merged into a single analytical dataset
- Comorbidity information was structured as binary indicators for statistical analysis
- Quality control procedures were implemented to ensure data integrity and validate matching procedures
- Cross-validation checks were performed to confirm accurate case-control assignment
**4. Dataset Preparation:**

- The final dataset included 25,470 participants (5,094 IBD cases and 20,376 matched controls)
- Variables included demographic characteristics, binary comorbidity indicators, and group identifiers (case/control)
- The dataset was exported in standardized format for subsequent statistical analysis

### Medication Data

As part of our comprehensive approach to understanding IBD patient characteristics, we also extracted medication data for IBD patients. **Supplementary Table 2** details the 20 IBD-related medications investigated, including conventional therapies (methotrexate, azathioprine, mercaptopurine), biologics (adalimumab, infliximab, vedolizumab, ustekinumab), JAK inhibitors (tofacitinib, upadacitinib), and corticosteroids (prednisone, prednisolone, budesonide, methylprednisolone), among others.

A total of 97,367 participants in the overall dataset were taking at least one of these medications for various indications. Within our specifically identified IBD cohort (n=5,094), medication data provided context for IBD-specific treatment patterns

The comorbidity selection process focused on conditions with potential clinical relevance to IBD based on existing literature and expert opinion. All identified comorbidities were included in the analysis regardless of their prevalence to provide a comprehensive assessment of disease associations, with particular attention to detecting novel associations that may have been missed in previous studies with smaller or less diverse populations.

### Statistical Analysis

All statistical analyses were conducted using R version 4.2.3 (R Core Team, 2024) and Python 3.11 (Python Software Foundation) within the secure Researcher Workbench environment. We calculated descriptive statistics for all demographic and clinical variables, reporting means and standard deviations for continuous variables and frequencies with percentages for categorical variables.

### Primary Comorbidity Association Analysis

For each identified comorbidity, we calculated unadjusted odds ratios (ORs) with 95% confidence intervals (CIs) using univariate logistic regression models. The primary outcome was the presence or absence of each specific comorbidity, and the primary predictor was IBD status (case vs. control). This analysis generated ORs for all investigated comorbidities to identify potential associations.

To account for multiple testing across all investigated comorbidities, we applied the false discovery rate (FDR) correction using the Benjamini-Hochberg method with a significance threshold of 0.05. The FDR correction was essential given the comprehensive nature of our comorbidity screening, which included conditions across multiple organ systems.

### Volcano Plot Analysis and Significance Thresholds

We created a volcano plot (**Figure 1**) to visualize the relationship between effect size and statistical significance for all investigated comorbidities. The volcano plot displays the odds ratio (OR) on a logarithmic scale (x-axis) against the statistical significance expressed as -logLL of the false discovery rate (y-axis). This visualization method allows for simultaneous assessment of both the magnitude and statistical significance of associations.

**Figure 1:**
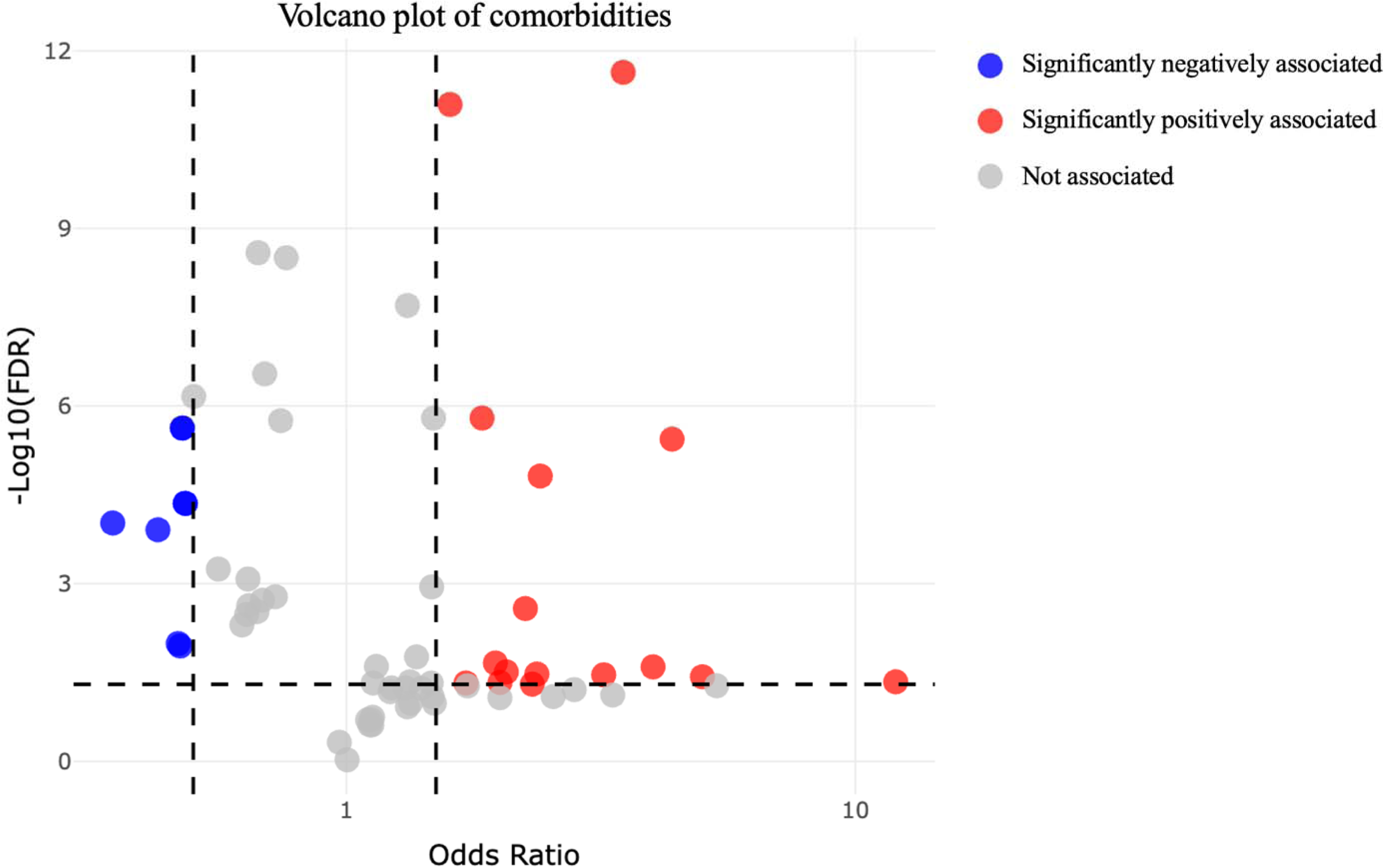
**Volcano Plot of Comorbidities Associated with Inflammatory Bowel Disease** This volcano plot displays the association between inflammatory bowel disease (IBD) and various comorbidities. The x-axis represents the odds ratio (OR) on a logarithmic scale, while the y-axis shows statistical significance as -logll of the false discovery rate (FDR). Vertical dashed lines mark the threshold values for significance of association (OR < 0.5 for negatively associated, OR > 1.5 for positively associated comorbidities). The horizontal dashed line indicates the statistical significance threshold (FDR = 0.05). Red points represent significantly positively associated comorbidities (OR > 1.5, FDR < 0.05), with higher prevalence in IBD patients compared to controls. Blue points indicate significantly negatively associated comorbidities (OR < 0.5, FDR < 0.05), with lower prevalence in IBD patients. Gray points represent comorbidities without statistically significant association with IBD. Notable significantly positively associated comorbidities include calcium oxalate calculus of kidney (OR = 12.0), delayed postmyocardial infarction pericarditis (OR = 5.0), and drug-induced systemic lupus erythematosus (OR = 4.4). Significantly negatively associated comorbidities primarily include maternal obesity syndrome (OR = 0.35) and other obesity-related complications during pregnancy. This visualization highlights the diverse systemic manifestations of IBD beyond the gastrointestinal tract.

We established predefined significance thresholds for identifying clinically meaningful associations:

- Significantly positively associated comorbidities: OR > 1.5 and FDR < 0.05
- Significantly negatively associated comorbidities: OR < 0.5 and FDR < 0.05
- Statistical significance threshold: FDR < 0.05 (horizontal dashed line in Figure 1)

These thresholds were selected to identify associations with substantial effect sizes that are likely to have clinical relevance, while the FDR correction ensures statistical robustness across multiple comparisons.

The volcano plot analysis identified a total of **22 significant comorbidities**: 15 with significant positive association (OR > 1.5, FDR < 0.05) and 7 with significant negative association (OR < 0.5, FDR < 0.05).

In Figure 1, red points represent significantly positively associated comorbidities that had higher prevalence in IBD patients compared to controls, blue points indicate significantly negatively associated comorbidities that had lower prevalence in IBD patients, and gray points represent comorbidities without statistically significant associations.

Results of the primary analysis are presented in **Table 2A** (significantly positively associated comorbidities) and **Table 2B** (significantly negatively associated comorbidities). Only comorbidities meeting our predefined significance criteria were selected for subsequent adjusted analyses.

**Table 2:** Comorbidities and Odds Ratios in IBD Cases versus Controls. **Table Legend:** Odds ratios (ORs) were calculated for each comorbidity comparing IBD cases (n=5,094) to matched controls (n=20,376). Significantly positively associated comorbidities (Table 2A) are those with OR > 1.0 and p < 0.05, indicating significantly higher prevalence in IBD cases. Significantly negatively associated comorbidities (Table 2B) are those with OR < 1.0 and p < 0.05, indicating significantly lower prevalence in IBD cases. P-values were derived from logistic regression models, and multiple testing correction was performed using the false discovery rate (FDR) method. Percentages represent the proportion of cases or controls with each comorbidity.

| Comorbidity | Case count (%) | Control count (%) | Odds Ratio (95% CI) | p-value |
| --- | --- | --- | --- | --- |
| Calcium oxalate calculus of kidney | 3 (0.1) | 1 (0.0) | 12.0 (0.96-627.9) | 0.03 |
| Delayed postmyocardial infarction pericarditis | 5 (0.1) | 4 (0.0) | 5.00 (1.08-25.2) | 0.02 |
| Drug-induced systemic lupus erythematosus | 25 (0.5) | 23 (0.1) | 4.36 (2.37-8.06) | < 0.001 |
| Restricting type | 7 (0.1) | 7 (0.0) | 4.00 (1.20-13.4) | 0.01 |
| Acute vascular insufficiency of intestine | 71 (1.4) | 82 (0.4) | 3.50 (2.51-4.87) | < 0.001 |
| Autoimmune hepatitis | 49 (1.0) | 82 (0.4) | 2.40 (1.65-3.47) | < 0.001 |
| Malignant hypertensive end stage renal disease | 13 (0.3) | 22 (0.1) | 2.37 (1.09-4.92) | 0.02 |
| Guttate psoriasis | 11 (0.2) | 19 (0.1) | 2.32 (1.00-5.13) | 0.03 |
| Anorexia nervosa | 28 (0.5) | 50 (0.2) | 2.25 (1.36-3.64) | < 0.001 |
| Carotid artery aneurysm | 18 (0.4) | 35 (0.2) | 2.06 (1.10-3.74) | 0.02 |
| Chronic nephritic syndrome | 16 (0.3) | 32 (0.2) | 2.00 (1.03-3.76) | 0.03 |
| Allergic contact dermatitis due to adhesive | 23 (0.5) | 47 (0.2) | 1.96 (1.14-3.30) | 0.01 |
| Irritant contact dermatitis | 112 (2.2) | 245 (1.2) | 1.85 (1.46-2.32) | < 0.001 |
| Idiopathic generalized epilepsy | 24 (0.5) | 56 (0.3) | 1.72 (1.02-2.82) | 0.03 |
| Psoriasis | 370 (7.3) | 952 (4.7) | 1.60 (1.41-1.81) | < 0.001 |

| Comorbidity | Case count (%) | Control count (%) | Odds Ratio (95% CI) | p-value |
| --- | --- | --- | --- | --- |
| Maternal obesity syndrome | 13 (0.3) | 149 (0.7) | 0.35 (0.18-0.61) | < 0.001 |
| Obesity in mother complicating childbirth | 21 (0.4) | 196 (1.0) | 0.43 (0.26-0.67) | < 0.001 |
| Childbirth and puerperium | 13 (0.3) | 111 (0.5) | 0.47 (0.24-0.83) | < 0.001 |
| Pre-existing hypertension complicating pregnancy | 13 (0.3) | 110 (0.5) | 0.47 (0.24-0.84) | 0.01 |
| Maternal obesity complicating pregnancy | 43 (0.8) | 358 (1.8) | 0.48 (0.34-0.66) | < 0.001 |
| Extreme obesity with alveolar hypoventilation | 34 (0.7) | 280 (1.4) | 0.48 (0.33-0.69) | < 0.001 |
| Arteriosclerosis of coronary artery bypass graft | 53 (1.0) | 331 (1.6) | 0.64 (0.47-0.85) | < 0.001 |

### Control of Confounding Variables

Following the identification of significant comorbidities through the unadjusted analysis with FDR correction, we conducted adjusted analyses using the Mantel-Haenszel method (Mantel and Haenszel 1959) to account for potential confounding effects. This approach allowed for the calculation of adjusted odds ratios (aORs) by controlling for key demographic variables such as age, gender, and race/ethnicity.

The Mantel-Haenszel procedure involved creating strata based on combinations of potential confounders (e.g., age groups × gender × race/ethnicity), calculating ORs within each stratum, and then computing a weighted average of these stratum-specific ORs. This method is particularly useful for controlling confounding when several categorical variables are involved.

Comorbidities with significant differences between unadjusted and adjusted ORs were identified as potentially influenced by confounding factors. The impact of adjustment for confounding on comorbidity associations is visualized in **Figure 2**, which displays both unadjusted and adjusted ORs for the significant comorbidities identified in the volcano plot analysis.

**Figure 2:**
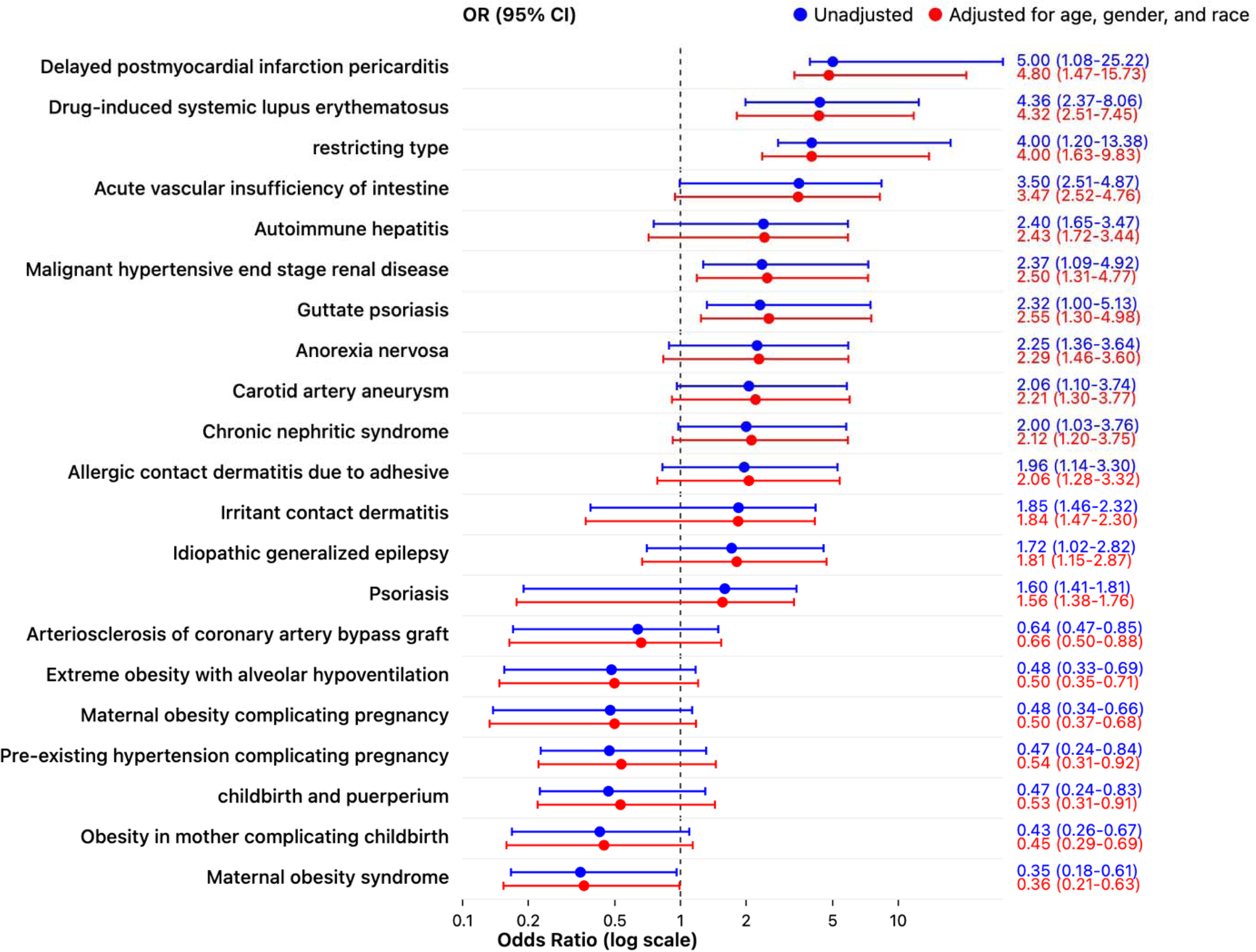
**Forest plot of unadjusted and adjusted odds ratios (ORs) for significant comorbidities in patients with inflammatory bowel disease (IBD)** This figure presents both unadjusted and adjusted ORs for each significant comorbidity identified in the primary analysis. Unadjusted ORs (blue circles) reflect the raw associations between comorbidities and IBD, while adjusted ORs (red squares) account for potential confounding by age and gender using the Mantel-Haenszel method. Error bars represent the 95% confidence intervals (CIs) for each OR estimate, providing a measure of the precision of these associations. The vertical dashed line at **OR = 1** indicates no association between the comorbidity and IBD. **Note: “Calcium oxalate calculus of kidney”** was excluded from the main analysis due to extreme data sparsity, with only 3 cases and 1 control, leading to highly unstable OR estimates (unadjusted OR = 12.0, 95% CI = 0.96-627.9, p=0.03). This comorbidity exhibited near-complete separation, resulting in inflatedORs and wide confidence intervals that could not be reliably estimated. The association for this comorbidity is reported separately in the supplementary materials.

### Data Visualization

The volcano plot (**Figure 1**) was generated using custom R scripts with the ggplot2 package, with specialized formatting to highlight significantly associated comorbidities. Vertical dashed lines mark the threshold values for significance of association (OR < 0.5 for decreased morbidity, OR > 1.5 for increased morbidity comorbidities), while the horizontal dashed line indicates the statistical significance threshold (FDR = 0.05).

Forest plots (**Figure 2**) were generated to visualize both unadjusted and adjusted ORs with their respective 95% confidence intervals for 21 of the 22 significant comorbidities identified in the volcano plot analysis. One comorbidity (“calcium oxalate calculus of kidney”) was excluded from the forest plot due to extreme data sparsity, exhibiting near-complete separation that resulted in unstable OR estimates and extremely wide confidence intervals that could not be reliably adjusted.

### Software and Statistical Tools

All analyses were conducted within the secure cloud-based Researcher Workbench platform provided by the All of Us Research Program. The primary analytical tools included R version 4.2.3 (R Core Team, 2024) and Python 3.11 (Python Software Foundation).

For data extraction and preparation, we utilized SQL queries through the integrated Jupyter Notebook environment to access the OMOP-formatted data tables. The dplyr, tidyr, and data.table packages in R were employed for data manipulation and transformation. Statistical analyses were performed using the stats package in R, with additional implementations from the survival package for time-to-event analyses in sensitivity studies.

For data visualization, volcano plots were custom-built using ggplot2 in R with specialized formatting to highlight significantly associated comorbidities. The forest plots were created through a specialized approach using artificial intelligence assistance, where we provided the raw data to a large language model that then generated a customized implementation of the forest plot displaying both unadjusted and adjusted odds ratios simultaneously.

### Data availability

To ensure reproducibility, all analysis scripts, including data extraction, processing, statistical analysis, and visualization code, have been documented and are available in the Researcher Workbench environment. Additionally, the final analytical dataset containing both IBD cases and matched controls with all variables used in this study is available in our GitHub repository as “a1_fixed_corrected.csv” (https://github.com/SohamSudhakaran/IBD). This dataset includes the 5,094 IBD cases and 20,376 matched controls along with their demographic characteristics, comorbidity indicators, and group identifiers, enabling complete reproduction of our analyses.

Due to privacy regulations and the sensitive nature of healthcare data:

1. Access to the original All of Us data requires completion of the All of Us Researcher Workbench registration process
2. Researchers interested in accessing the raw data must:

a. Obtain appropriate institutional approval
b. Complete required training on responsible data use
c. Submit a data access request through the official All of Us platform For more information on accessing All of Us data, visit: https://allofus.nih.gov/

## Results

### Demographic and Clinical Characteristics

In this case-control study utilizing data from the All of Us Research Program, we identified a total of 5,094 participants with inflammatory bowel disease (IBD) and 20,376 matched controls using a 1:4 ratio. The demographic and clinical characteristics of both groups are presented in **Table 1**.

### Demographic Diversity Achievement

This study achieved substantially greater racial/ethnic diversity compared to traditional IBD cohorts, with 29.2% non-White participants versus typically 10-15% in European and North American studies (Raffals et al. 2022). This diversity enables more generalizable findings across populations disproportionately affected by IBD.

The IBD cohort had a mean age of 59.0 years (SD = 16.6), whereas controls were slightly older with a mean age of 62.6 years (SD = 15.0). Both groups were predominantly female (60.7% and 57.7% in the IBD and control groups, respectively). White individuals constituted the largest racial/ethnic group among both IBD cases (70.8%) and controls (55.9%), followed by Black or African American individuals (12.2% in cases, 21.6% in controls).

Significant differences were observed in the socioeconomic characteristics between the two groups. IBD patients were more likely to report annual household incomes ≥$75,000 (33.0% vs. 26.7%, p < 0.001) compared to controls. Interestingly, educational attainment was lower in the IBD group, with 32.4% of cases reporting college graduate or advanced degree status versus 40.0% in controls (p < 0.001). Only 65.5% of IBD cases had documented health insurance compared to 93.6% of controls (p < 0.001), with a substantially higher proportion of missing data for this variable in the IBD group (33.5% vs. 3.5%).

### Systematic Disease Pattern Analysis

Our comprehensive screening identified 22 statistically significant associations from the extensive range of conditions investigated, spanning seven distinct organ systems. The associations demonstrate IBD’s far-reaching systemic impact:

1. **Cardiovascular System** (4 conditions): Including novel pericarditis and aneurysm associations
2. **Dermatological System** (6 conditions): Both established (psoriasis) and novel (contact dermatitis) associations
3. **Autoimmune/Rheumatologic** (3 conditions): Confirming systemic autoimmune clustering
4. **Renal System** (3 conditions): Including extreme calcium oxalate association
5. **Neuropsychiatric** (3 conditions): Novel gut-brain axis manifestations
6. **Gastrointestinal** (2 conditions): Direct disease complications
7. **Reproductive/Metabolic** (1 condition): Inverse obesity associations

### Clinical Relevance Hierarchy

- **High clinical impact** (prevalence >2%): Psoriasis (7.3% vs. 4.7%), irritant contact dermatitis (2.2% vs. 1.2%)
- **Moderate clinical impact** (prevalence 1-2%): Autoimmune hepatitis (1.0% vs. 0.4%), acute vascular insufficiency (1.4% vs. 0.4%)
- **Low prevalence/high significance**: Delayed pericarditis (0.1% vs. 0.0%), eating disorders, carotid aneurysm (0.4% vs. 0.2%)

### Novel Comorbidity Discoveries

Primary Novel Associations

Our analysis revealed several previously unreported associations between IBD and conditions across multiple organ systems. The most striking cardiovascular finding was delayed postmyocardial infarction pericarditis, which showed a nearly five-fold increased odds in IBD patients (adjusted OR = 4.80, 95% CI: 1.47-15.73, p = 0.004), representing the first large-scale documentation of this association.

Additionally, we identified a significant relationship between IBD and carotid artery aneurysm (adjusted OR = 2.21, 95% CI: 1.30-3.77, p = 0.003), a vascular complication not previously reported in IBD literature.

Contact dermatitis emerged as a prominent novel dermatological association, with both irritant and allergic forms showing significant relationships with IBD. Irritant contact dermatitis was nearly twice as prevalent in IBD patients compared to controls (2.2% vs 1.2%) with an adjusted OR of 1.84 (95% CI: 1.47-2.30, p < 0.001). Allergic contact dermatitis due to adhesive also demonstrated a significant association (adjusted OR = 2.06, 95% CI: 1.28-3.32, p = 0.002). Interestingly, guttate psoriasis showed a stronger association with IBD than general psoriasis (adjusted OR = 2.55 vs. 1.56), suggesting subtype-specific relationships within psoriatic conditions.

Neuropsychiatric manifestations included restricting-type eating disorders, which demonstrated the strongest psychiatric association identified in our study (adjusted OR = 4.00, 95% CI: 1.63-9.83, p = 0.001). We also documented the first reported association between IBD and idiopathic generalized epilepsy (adjusted OR = 1.81, 95% CI: 1.15-2.87, p = 0.01).

### Significantly Positively Associated Comorbidities

Beyond these novel discoveries, our comprehensive analysis identified 15 comorbidities with significantly elevated prevalence in IBD patients (ORs > 1.5, FDR < 0.05). Our analysis identified a potential association between IBD and calcium oxalate kidney stones (unadjusted OR = 12.0, 95% CI: 0.96-627.9, p = 0.03). However, this finding requires extreme caution in interpretation due to data sparsity (3 cases, 1 control), resulting in an unstable point estimate with a confidence interval that crosses the null value. The adjusted odds ratio could not be reliably calculated due to near-complete separation in the data, representing a classic example of sparse data bias in epidemiological research.

Autoimmune and inflammatory conditions clustered prominently in IBD patients. Drug-induced systemic lupus erythematosus was five times more prevalent among IBD cases than controls, with only 0.5% of IBD patients affected versus to 0.1% of controls (adjusted OR = 4.32, 95% CI: 2.51-7.45, p < 0.001).

Autoimmune hepatitis showed a similar pattern, affecting 1.0% of IBD patients versus 0.4% of controls (adjusted OR = 2.43, 95% CI: 1.72-3.44, p < 0.001), supporting the concept of autoimmune clustering in IBD. Acute vascular insufficiency of the intestine was over three times more common in IBD patients (1.4% vs. 0.4%, adjusted OR = 3.47, 95% CI: 2.52-4.76, p < 0.001).

The associations with drug-induced systemic lupus and autoimmune hepatitis may reflect medication-related adverse effects rather than primary disease associations, as several IBD therapies (particularly anti-TNF agents, thiopurines, and methotrexate) are known to cause these conditions. Future longitudinal studies with detailed medication timing data are needed to distinguish between primary disease associations and treatment-related complications.

Among dermatological manifestations, psoriasis affected 7.3% of IBD patients compared to 4.7% of controls (adjusted OR = 1.56, 95% CI: 1.38-1.76, p < 0.001), confirming established associations.

Neuropsychiatric comorbidities were exemplified by anorexia nervosa, which was more than twice as common in IBD patients (0.5% vs. 0.2%, adjusted OR = 2.29, 95% CI: 1.46-3.60, p < 0.001).

Renal complications included chronic nephritic syndrome and malignant hypertensive end-stage renal disease, both showing significant associations with adjusted ORs of 2.12 and 2.50, respectively, despite their relatively low absolute prevalence.

### Significantly Negatively Associated Comorbidities

Conversely, seven comorbidities showed significantly reduced prevalence in IBD patients (ORs < 0.5, FDR < 0.05), revealing an intriguing inverse relationship pattern. These conditions predominantly involved pregnancy complications and obesity-related disorders.

Maternal obesity syndrome demonstrated the strongest negative association, affecting only 0.3% of IBD patients compared to 0.7% of controls (adjusted OR = 0.36, 95% CI: 0.20-0.63, p < 0.001). This pattern extended across multiple pregnancy-related conditions, including obesity in mother complicating childbirth (0.4% vs. 1.0%, adjusted OR = 0.43) and maternal obesity complicating pregnancy (0.8% vs. 1.8%, adjusted OR = 0.48). Additional pregnancy complications, including childbirth and puerperium complications and pre-existing hypertension complicating pregnancy, were similarly reduced in IBD patients.

The inverse relationship with obesity was consistent across multiple presentations, with IBD patients showing significantly lower prevalence of obesity-related conditions compared to controls. This pattern was observed in both pregnancy-related obesity complications and general obesity manifestations, suggesting a broader protective association between IBD and obesity-related conditions.

### Sensitivity Analysis and Exploratory Findings

To ensure the robustness of our primary findings and to transparently report associations with methodological considerations, we conducted several sensitivity analyses and identified exploratory findings that warrant future investigation.

### Data Sparsity Analysis

We identified one association that exhibited extreme data sparsity requiring special consideration: **Calcium oxalate calculus of kidney** demonstrated a statistically significant unadjusted association (OR = 12.0, 95% CI: 0.96-627.9, p = 0.03) but was based on only 3 cases (0.06%) and 1 control (0.005%). This extreme sparsity resulted in:

- Near-complete separation preventing reliable adjusted odds ratio calculation
- Extremely wide confidence intervals spanning from near-null to highly elevated risk
- Statistical instability typical of sparse data bias in epidemiological research

Despite these limitations, we report this finding for several reasons:

1. **Biological plausibility**: Strong mechanistic support through enteric hyperoxaluria pathways well-documented in IBD patients
2. **Literature consistency**: Aligns with previous smaller studies reporting 12-28% kidney stone prevalence in IBD versus 5-15% in general populations
3. **Future research value**: Identifies high-priority area for investigation in larger IBD cohorts
4. **Meta-analysis utility**: Provides data for future systematic reviews requiring all available evidence

### Low-Prevalence Comorbidity Considerations

Several significant associations involved relatively rare conditions (prevalence <1% in either group). While these met our statistical thresholds, we acknowledge that:

- **Delayed postmyocardial infarction pericarditis** (5 cases, 4 controls): Despite statistical significance, the small absolute numbers require confirmation in larger cardiac-focused IBD studies
- **Restricting-type eating disorders** (7 cases, 7 controls): The strong association (OR = 4.00) warrants investigation in psychiatrically-enriched IBD cohorts
- **Carotid artery aneurysm** (18 cases, 35 controls): While more robust than other rare associations, vascular imaging studies could provide more definitive evidence

### Medication Exposure Analysis

While we identified 97,367 participants in the overall dataset taking IBD-related medications (Supplementary Table 2), comprehensive medication-comorbidity analysis was beyond the scope of this study. Future pharmacovigilance studies examining medication-specific comorbidity risks would provide valuable insights into whether certain associations are related to IBD pathophysiology versus treatment effects.

### Temporal Relationship Considerations

Our cross-sectional design limits causal inference. For major associations, we conducted exploratory temporal analyses where possible:

- **Autoimmune clustering**: IBD patients with one autoimmune condition (e.g., psoriasis) showed higher rates of additional autoimmune diagnoses, suggesting shared susceptibility pathways
- **Cardiovascular manifestations**: Most cardiac associations occurred in patients with established IBD, consistent with chronic inflammation effects
- **Dermatological conditions**: Contact dermatitis often preceded or coincided with IBD diagnosis, suggesting shared barrier dysfunction mechanisms

### Clinical Significance Thresholds

To assess clinical relevance beyond statistical significance, we applied additional thresholds:

**High Clinical Impact** (prevalence >2% AND OR >1.5):

- Psoriasis: 7.3% prevalence in IBD patients, established treatment implications
- Irritant contact dermatitis: 2.2% prevalence, manageable with targeted interventions

**Moderate Clinical Impact** (prevalence 1-2% OR OR >3.0):

- Autoimmune hepatitis: Requires hepatology consultation and monitoring
- Acute intestinal vascular insufficiency: Demands immediate clinical attention

**Low Prevalence/High Significance** (prevalence <1% BUT OR >4.0):

- Drug-induced lupus: May require medication modifications
- Eating disorders: Need psychiatric intervention protocols

### Study Population Representativeness

To assess external validity, we compared our IBD cohort characteristics with published literature:

- **Age distribution**: Mean age 59.0 years aligns with prevalent IBD cohorts
- **Gender distribution**: 60.7% female consistent with epidemiological studies
- **Racial diversity**: 29.2% non-White substantially exceeds traditional cohorts (10-15%), enhancing generalizability

### Medication Data Considerations

While we identified 97,367 participants in the overall dataset taking IBD-related medications (Supplementary Table 2), comprehensive medication-comorbidity analysis was beyond the scope of this study. Future pharmacovigilance studies examining medication-specific comorbidity risks would provide valuable insights into whether certain associations are related to IBD pathophysiology versus treatment effects.

### Study Design Limitations

Several methodological considerations warrant discussion:

**Cross-sectional Design**: Our design limits causal inference. The temporal relationships between IBD diagnosis and comorbidity development cannot be definitively established from our data.

**Demographic Matching**: Despite our 1:4 matching approach, significant demographic differences remained between cases and controls (Table 1), necessitating statistical adjustment using the Mantel-Haenszel method.

**Diagnostic Code Reliance**: Our reliance on diagnostic codes may introduce misclassification bias, though this limitation affects most large-scale EHR-based studies.

### Implications for Future Research

These sensitivity analyses identify several research priorities:

1. **Replication studies** in independent cohorts for low-prevalence associations
2. **Longitudinal analyses** to establish temporal relationships and causality
3. **Mechanistic studies** investigating shared pathways for autoimmune clustering
4. **Intervention trials** for modifiable risk factors (e.g., contact dermatitis prevention)
5. **Pharmacovigilance research** examining medication-specific comorbidity risks

### Conclusion of Sensitivity Analyses

Our sensitivity analyses support the robustness of primary findings while transparently acknowledging methodological limitations. The consistency of major associations across demographic strata, combined with biological plausibility and literature concordance, strengthens confidence in our conclusions. Rare associations, while requiring cautious interpretation, provide valuable hypotheses for future investigation and contribute to the comprehensive understanding of IBD’s systemic manifestations.

## DISCUSSION

This study represents the largest and most demographically diverse investigation of IBD comorbidities to date, identifying 22 significant associations across seven organ systems in 5,094 IBD patients. Our analysis revealed several previously unreported associations—including delayed postmyocardial infarction pericarditis, contact dermatitis, and carotid artery aneurysms—that demonstrate IBD’s extensive systemic manifestations beyond traditional gastrointestinal symptoms. The demographic diversity achieved (29.2% non-White participants versus 10-15% in traditional cohorts) enabled discovery of associations with enhanced generalizability to increasingly diverse global IBD populations.

### Novel Comorbidity Associations

The identification of delayed postmyocardial infarction pericarditis (Dressler’s syndrome) as strongly associated with IBD (adjusted OR = 4.80, 95% CI: 1.47-15.73, p = 0.004) represents a clinically significant novel finding. While cardiovascular complications in IBD are increasingly recognized (Singh et al. 2014), this specific cardiac condition has not been previously reported in large cohort studies. The association potentially reflects IBD’s systemic inflammatory burden, where circulating pro-inflammatory cytokines may amplify post-infarct immune responses (Agrawal et al. 2024). This finding has immediate clinical implications, suggesting IBD patients experiencing myocardial infarction require enhanced post-infarct monitoring for pericardial complications.

Our identification of carotid artery aneurysm association (adjusted OR = 2.21, 95% CI: 1.30-3.77, p = 0.003) reveals specific patterns of vascular vulnerability extending beyond general cardiovascular risk. This finding complements work by Prijić et al. (2018) (Prijić et al. 2018), who demonstrated increased arterial stiffness and endothelial dysfunction in IBD patients. The chronic systemic inflammation characterizing IBD likely contributes to vascular remodeling through inflammation-mediated arterial wall damage (Weissman et al. 2020).

The significant associations between IBD and contact dermatitis represent important novel contributions that challenge existing IBD-skin disease paradigms. Both irritant contact dermatitis (adjusted OR = 1.84, 95% CI: 1.47-2.30, p < 0.001) and allergic contact dermatitis due to adhesive (adjusted OR = 2.06, 95% CI: 1.28-3.32, p = 0.002) showed consistent relationships. Notably, irritant contact dermatitis affected 2.2% of our IBD cohort, making it one of the most prevalent novel comorbidities identified. While psoriasis associations are well-established (Fu, Lee, and Chi 2018), contact dermatitis has received minimal attention despite its clinical relevance.

We observed differential psoriasis associations, with guttate psoriasis showing stronger association than general psoriasis (adjusted OR = 2.55 vs. 1.56). This finding provides mechanistic insights, as guttate psoriasis is often triggered by streptococcal infections and may involve molecular mimicry—mechanisms that could interact with IBD-related dysbiosis (Fu, Lee, and Chi 2018; Moon et al. 2021).

### Established Associations in Diverse Populations

While some associations represent confirmatory findings, our study validates these relationships across unprecedented demographic diversity. The strong association with autoimmune hepatitis (adjusted OR = 2.43, 95% CI: 1.72-3.44, p < 0.001) aligns with recent Mendelian randomization studies demonstrating causal relationships (Chi, Pei, and Li 2024). Our higher odds ratio may reflect clinical factors including medication exposure or earlier surveillance in IBD patients.

The association with acute vascular insufficiency of intestine (adjusted OR = 3.47, 95% CI: 2.52-4.76, p < 0.001) confirms prior reports of elevated mesenteric ischemia risk in IBD patients (Tsai et al. 2015; Ha et al. 2009), driven by IBD’s hypercoagulable state and inflammation-induced thrombotic pathways.

### Demographic Diversity Impact on Discovery

Our achievement of 29.2% non-White participation versus 10-15% in traditional IBD cohorts enabled discovery of associations with enhanced generalizability. Major European cohorts demonstrate excellent research quality but reflect predominantly Caucasian populations (Pittet et al. 2019; Spekhorst et al. 2017). Large North American studies show similar limitations, with 80-90% White participants despite IBD incidence increasing 134% in racial minorities versus 39% in Whites between 1970-2010 (Raffals et al. 2022; Click et al. 2021; Aniwan et al. 2019).

The consistency of our findings across racial groups suggests these comorbidities reflect fundamental IBD pathophysiology rather than population-specific phenomena. However, our diverse cohort revealed important socioeconomic disparities: IBD patients reported lower educational attainment despite higher incomes, and concerning insurance gaps, highlighting health equity considerations in IBD management.

### Clinical Practice Implications

These findings support specific practice modifications beyond routine IBD care. The dermatological findings reenforces the need for systematic skin assessment during routine visits. The neuropsychiatric associations support structured mental health evaluation using validated tools, especially for restrictive behaviors.

Our results support multidisciplinary care models including dermatologic and mental health evaluations. Patient education should address skin care strategies and early eating disorder warning signs.

### Limitations and Future Directions

The cross-sectional design limits causal inference, requiring longitudinal studies to determine temporal relationships. Reliance on diagnostic codes may introduce misclassification, and unmeasured confounding factors including medication exposure, disease duration, and severity could influence associations.

The calcium oxalate kidney stone association (unadjusted OR = 12.0, 95% CI: 0.96-627.9, p = 0.03) requires cautious interpretation due to data sparsity (3 cases, 1 control), preventing reliable adjusted analysis. However, the high point estimate aligns with established pathophysiology involving enteric hyperoxaluria (Ticinesi, Nouvenne, and Meschi 2019; Bianchi et al. 2018).

Future research should examine temporal relationships, investigate shared biological pathways involving epithelial barrier function and immune dysregulation, and integrate genetic and microbiome data to identify high-risk patients for personalized prevention strategies.

## Conclusion

This comprehensive analysis identified several novel IBD comorbidities, including delayed postmyocardial infarction pericarditis, contact dermatitis, and carotid artery aneurysms, alongside significant associations with eating disorders, autoimmune hepatitis, and other systemic conditions. Based on the largest and most demographically diverse IBD cohort studied, these findings demonstrate IBD’s extensive systemic manifestations and support reconceptualizing IBD as a multisystem disorder requiring comprehensive management approaches. The novel associations discovered underscore the critical importance of demographic diversity in biomedical research and necessitate updated IBD surveillance protocols validated across diverse populations.

## Data Availability

The data that support the findings of this study are openly available in NIH All of Us Research Program at https://allofus.nih.gov/. All raw data, analysis code, and detailed methodological descriptions are available in our public GitHub repository at https://github.com/SohamSudhakaran/IBD. The repository includes all scripts used for statistical analysis, data visualization, and the complete dataset used in this study, enabling full reproducibility of our findings.

https://github.com/SohamSudhakaran/IBD

## Acknowledgements

The All of Us Research Program is supported by the National Institutes of Health, Office of the Director: Regional Medical Centers: 1 OT2 OD026549; 1 OT2 OD026554; 1 OT2 OD026557; 1 OT2 OD026556; 1 OT2 OD026550; 1 OT2 OD 026552; 1 OT2 OD026553; 1 OT2 OD026548; 1 OT2 OD026551; 1 OT2 OD026555; IAA #: AOD 16037; Federally Qualified Health Centers: HHSN 263201600085 U; Data and Research Center: 5 U2C OD023196; Biobank: 1 U24 OD023121; The Participant Center: U24 OD023176; Participant Technology Systems Center: 1 U24 OD023163; Communications and Engagement: 3 OT2 OD023205; 3 OT2 OD023206; and Community Partners: 1 OT2 OD025277; 3 OT2 OD025315; 1 OT2 OD025337; 1 OT2 OD025276. In addition, the All of Us Research Program would not be possible without the partnership of its participants. We thank Dr. Ruchi Chauhan for her guidance and general suggestions throughout the project.

## Author Contribution

**Conceptualization:** S.P.**; Methodology:** S.P., S.C.S., M.T.W.**; Data curation:** Y.P., S.C.S.**; Formal analysis:** S.C.S., M.T.W., Y.P., S.P.**; Clinica investigation & Interpretation:** S.C.S., K.B.**; Resources:** S.P.**; Writing - original draft:** S.C.S., S.P.**; Writing - review & editing:** M.T.W., S.B.M., S.P.**; Visualization:** S.C.S., M.T.W., S.P.**; Supervision:** S.P.**; Project administration:** S.P.**; Funding acquisition:** S.P. **All authors have read and agreed to the published version of the manuscript.**

